# Impact of spatial aggregation on detection of spatiotemporal disease clusters: analysis of SARS-CoV-2 infections in 3-D high-density settings

**DOI:** 10.1101/2024.12.20.24318345

**Authors:** Keith Allison, Andrew A. Lover

## Abstract

**Introduction:** High-density congregate housing, including cruise ships, hotels, residence halls and correctional facilities are epidemiologically important, and key aspects of pathogen transmission have been elucidated in these environments. A range of methods have been developed to detect unusual clusters of infections in these settings; however use of explicitly 3-D (x,y,z) spatial data has received little attention. In this study, we use data collected during the COVID-19 pandemic to assess the fine-scale spatial epidemiology and the clustering of confirmed cases to better understand impacts of spatial resolution and aggregation on spatio-temporal cluster detection.

**Methods:** Data for this analysis combined the results from mandatory weekly viral testing during the 2020-2021 academic year with high-resolution spatial data from university students residing in high-rise residence halls at the University of Massachusetts, Amherst campus.

These data were analyzed for statistically-significant clustering of SARS-CoV-2 cases in three-dimensional space as well as time, within and between the high-density buildings on campus. Two sets of analyses were conducted. The first used a Space-Time Permutation Model, which scans for areas with a greater than expected number of cases (SaTScan). To assess the impact of data aggregation, analysis was done at several levels of spatial resolution. Additionally, we performed sensitivity analyses using a purely temporal surveillance algorithm, CDC’s Early Aberration Reporting System-EARS.

**Results and conclusions:** Analysis with SaTScan at the room- and floor-level identified multiple statistically significant clusters within one residence hall. Analyses with these same cases aggregated at the floor-level were found to be as sensitive, but far less computationally intensive, than room-level analysis. Analysis at both of these spatial scales was more sensitive than analysis aggregated at the street address-level. Two events exceeding alert thresholds were detected in the purely temporal analysis; one of which was also detected in spatio-temporal analyses.

These results expand our understanding of spatio-temporal scan metrics to include 3-D analysis, and optimizing choice of spatial scales. These results have broad applicability in epidemiology in assessing the ability of spatio-temporal methods for public health surveillance, with potential expansion to ecological studies incorporating vertical movement.

## 1 Introduction

Health surveillance is a core activity of public health; and detection of anomalous case counts is critical for monitoring and response. The first cases of novel coronavirus disease 2019 (COVID-19) appeared in Wuhan City, China in December of 2019 [1]. This virus quickly spread globally, with the WHO declaring a PHEIC (Public Health Emergency of International Concern) in January 2020 and a pandemic in March 2020. Public health organizations rapidly began gathering, analyzing, and disseminating data about the spread of SARS-CoV-2. In the United States, major efforts were rapidly implemented to track and forecast the spread of the virus, including using spatial, temporal, and spatial-temporal methods to detect potential clusters of cases. SARS-CoV-2 infections are an ideal testing-ground for these methods due to massive-scale testing, high-resolution data, and well-understood natural history of infections across the subsequent pandemic waves.

### 1.1 Case-cluster detection methods and aggregation

Many outbreak detection algorithms are purely temporal and these are the most widely used in public health surveillance [2]; however, spatial-temporal methods are increasingly used for epidemiological surveillance and research.

There are multiple scenarios for use of these expanded methods. For example, Kan et al used spatial-temporal methods to identify high risk areas in Hong Kong with elevated numbers of reported COVID-19 infections. They suggest that their research can inform policy, lead to more targeted prevention strategies, and impact the behavior of the public. Dellicour et al also used spatial-temporal methods to determine factors associated with COVID-19 hospitalization to predict future hospitalizations[3].

A large field of research exists developing “optimal” methods for outbreak detection using spatial-temporal methods. Deeper understanding of the spatial-temporal nature of outbreaks has the potential to allow for more efficient and effective responses in the future. The spread of communicable diseases in both space and time has been extensively studied, especially in the context of emerging diseases like COVID-19. The Space-time Scan Statistic (STSS), developed by Kulldorf and collaborators, combines both the spatial and temporal elements of disease analysis to detect clusters, and has been used both prospectively, to predict new clusters of COVID-19, and retrospectively, to analyze previously collected data for clusters.

*Retrospective* analysis of outbreak infection data is important for optimizing outbreak detection systems, exploring alternative detection methods, and identifying areas of concern or previously undetected clusters to understand underlying epidemiology. A “perfect” outbreak detection system should identify all outbreaks at a given threshold, while giving minimal false positives. Of course, this requires adjustment and testing of algorithm parameters. Retrospective analysis provides a suitable environment for such testing, as sensitivity can be adjusted to detect outbreaks that were identified by active surveillance and vetted before being implemented for prospective surveillance.

Data aggregation has been explored in several studies; many studies are analyzed at the post code or county-level. For exmaple, Ling et al studied dengue fever in Malaysia at both the street address and sub-district (county) level and explored the effects of spatial scale on analysis[4]. This work suggests that incorporation of both spatial and temporal elements of dengue transmission were needed to support control measures.

### 1.2 3-D data in high-density living environments

There has been limited use of 3-D spatial data in epidemiological analysis, likely due to the difficulties in obtaining exact (x,y,z) coordinates. Studies in high-density settings like worker dormitories in Singapore looked at spatial arrangements, and concluded that the building design and layout may play an important role, but did not quantify the spatial aspects of the epidemiology [5]. Another other study in Hong Kong did examine overall building height as a “risk factor” (high-rise versus low-rise) for SARS-CoV-2, but did not assess spatial clustering of cases within this context. [6].

A recent review of spatio-temporal methods for outbreak detection [7] highlights a dearth of studies using 3D plots to depict temporal trends in x-y coordinate data, and while several ecological studies have considered the impacts of vertical movements [8, 9] in population dynamics, we are not aware of any quantitative “hotspot” analysis in 3-D systems.

### 1.3 Study rationale

In response to the pandemic, the University of Massachusetts, Amherst (hereafter, UMass) suspended all in-person classes on March 13th, 2020, and on March 20th, 2020 finalized plans to relocate all remaining students living on campus. The campus continued to be empty of students for the remainder of that academic year. During the subsequent academic year (2020-2021) students returned to campus with measures in place to mitigate the spread of the virus. Masking was required in all indoor areas, and for a period of time outdoors as well. In-class learning was minimal (essential lab-based courses only) and most learning was done remotely. Students living on-campus as well as students who did not live on campus but attended in-person classes were required to submit to frequent (weekly, or bi-weekly) testing. Those students who tested positive were moved to isolation residences and interviewed by staff from the University’s Public Health Promotion Center (PHPC). This subsequent contact tracing also recorded reported symptoms, and geographic locations. These interview data were captured into a centralized HIPAA-compliant database for analysis.

These data collected by the PHPC had a high spatial resolution (including both floor and room numbers) compared to data generally collected for outbreak detection purposes. Cluster detection methods generally use data at larger spatial scales (county-level, or postal code), or by neighborhood. Well-georeferenced data in high-density living environments are important epidemiological tools, and due to institutional record keeping often provide critical epidemiological insights that would not be possible using routine household level data. Even event detection done at the street- or address-level lacks information about floor and room location in three-dimensional space. However, exact location data requires specific workflows and data collection as well as building floor plans and complex and time-consuming geolocation that cannot currently be done by address geolocation APIs.

However, STSS is capable of cluster detection at finer spatial scales. This could be at the address level or even down to the room level, in the case of apartment buildings or dormitories. This level of resolution can help detect the localized spread of disease and has been done by Abboud et al to study the spread of *Klebsiella pneumoniae* in a single, high complexity hospital[10]. Our goal is to assess the implementation of STSS at this high-resolution, room-specific, level and determine if significant clustering occurs within dormitory buildings, and to assess sensitivity in comparison to coarser spatial scales.

## 2 Methods

All analyses were performed using R software [11]. Other packages included **RSatScan** [12],, **plotly** [13], and **surveillance** [14]. Statistical significance throughout was at the *α* = 0.05 level.

### 2.1 Study population

We utilized data collected by the UMass Public Health Promotion Center (PHPC) for laboratory-confirmed SARS-CoV-2 infections (PCR-based, in a CLIA-certified facility). At the time of the study (Spring 2021), the university had mandatory weekly asymptomatic testing for all students and faculty/staff, and the on-campus CLIA lab (Institute for Applied Life Sciences) was performing approximately 5,000 tests per day.

Data for all positive lab testing were followed up by contact-tracers, and relevant demographic details collected. We extracted all confirmed cases from three large residential facilities (Dorms J, K, and C) from February 1st, 2021, to May 15th 2021. These data included for each individual the date of infected sample collection, the date of symptom onset, the dormitory building, gender, floor number, and room number. The presumed date of infection was either the date of sample collection or reported symptom onset date, whichever was earlier. Figure 1 shows total reported cases by date across each of the three residence halls. During this time period there was an obvious uptick in incidence across campus; some individual residence halls not included in this analysis reported >10 new cases per 100 population per week.

**Figure 1.**
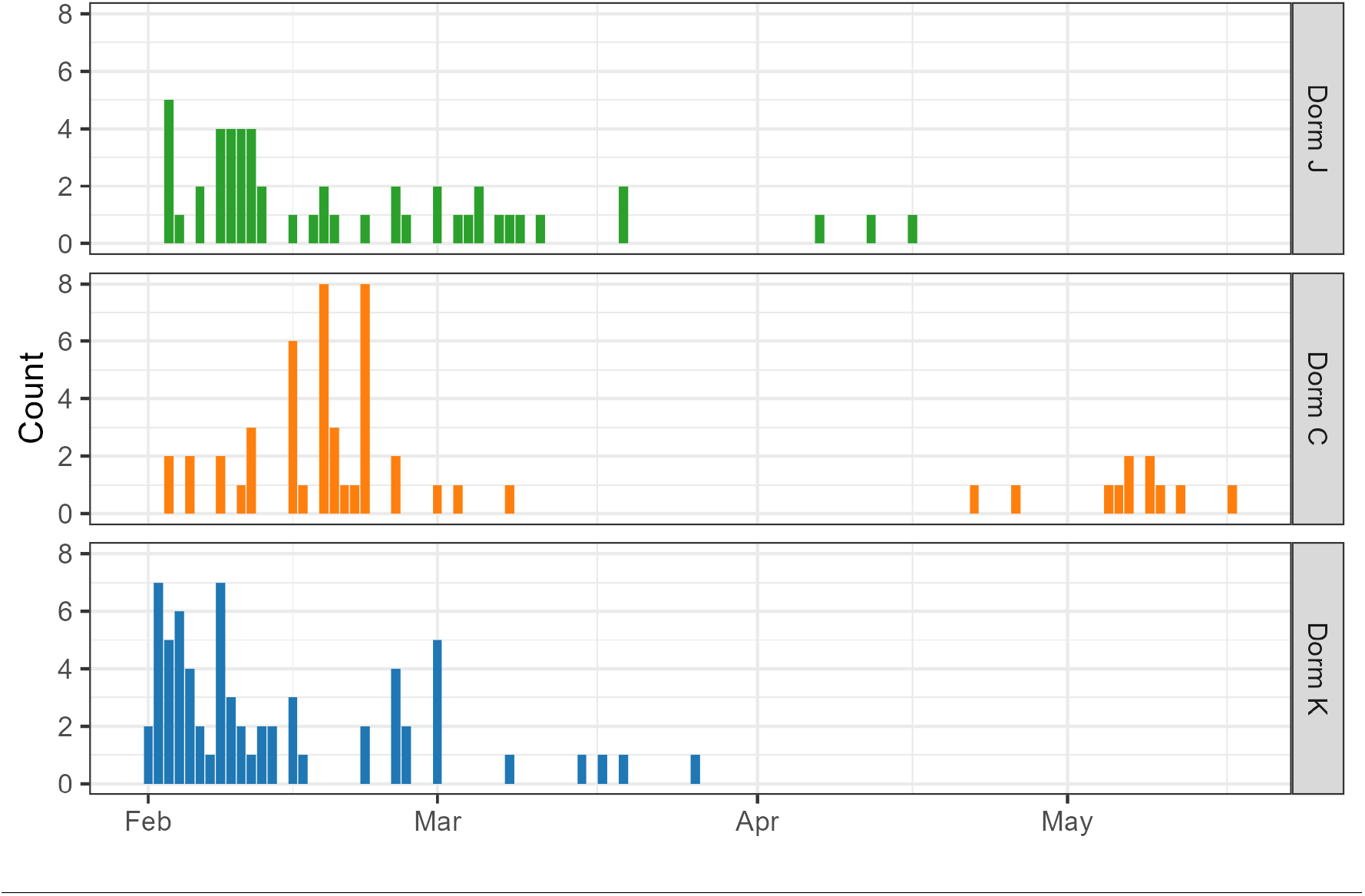
SARS-CoV-2 case counts by residence hall building, 2021.

All data were collected as part of routine campus public health response, which was reviewed by University of Massachusetts-Amherst IRB, and determined to not be human subjects research (UMass HRPO reference: #20-258). A separate protocol was submitted for this analysis, and was also determined to not be human subjects research (UMass HRPO determination: #6221).

### 2.2 Spatial data sources

We used QGIS (version 3.22.3[15]) to overlay floor plans obtained from the university residential services office onto building outlines (latitude-longitude), referenced using Google earth. We determined the location of each room by geolocating the center point. The elevation of each room was estimated using the total height of the buildings (in meters), and assuming evenly-spaced floors. Together, these data allowed us to locate the relative position of each reported case in three-dimensional space. Each residence hall has three elevators, and two stairwells all located on one end of the building. The distances between cases were determined in a Euclidean fashion; that is, in direct lines between cases in space.

### 2.3 Space-Time Permutation Scan Statistic

We used the retrospective Space-Time permutation scan statistic contained in the SaTScan program v. 10.0.1, [16]) to retrospectively detect clusters of SARS-CoV-2 infections that appeared during the time period of interest. This method requires no controls or background population data-instead it compares the number of observed cases in a cluster with the expected cases in a cluster, given that there was no space-time interaction (SaTScan guide [16]). That is, this model detects geographic areas with a higher-than-expected number of cases in a certain time period compared to other geographic areas at that time.

In a two-dimensional space, SaTScan uses a cylinder as the scanning window, with the circular base representing the geography of the cases and the height of the cylinder representing the time period [17]. In three-dimensional space, the scanning window is a sphere, with time projected into a fourth dimension. Each scanning window then is centered on each case, and the temporal and spatial window is adjusted and iterated until it reaches set boundaries.

In this study, the temporal boundaries were a minimum of one day to a maximum of 50% of the total study period. The spatial boundaries were restricted to at most 50% of the population-at-risk, and cluster were required to have at least two cases to be flagged. The scan statistic was also adjusted for the covariates of patient gender and day the week to adjust for potential testing and reporting biases. We conducted the analysis on each residence hall individually, as well as all three buildings together in aggregate.

The space-time permutation scan statistic was proposed and further described [18]. As with other scan statistics, the space-time permutation model detects areas in space and time of unexpectedly high case counts. However, unlike other scan statistics it does not require a uniform population at risk, a control group, or other baseline information. It also differs from other scan statistics by its probability model; the expected number of cases is determined using only the cases. In this case C is the number of observed cases and *c*_*zd*_ is the number of cases in area z during day d.

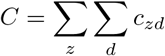

For each area and day, we calculated the expected number of cases *µ*_*zd*_ conditioning on the observed marginals.

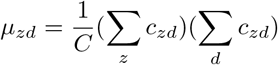

This value is the proportion of all cases in area z times the total cases during day d. *µ*_*A*_ is the expected number of cases inside the scanning window A.

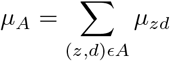

We assume that the probability of a case being in an area z given that it was observed on day d is the same for all days d. When there is no space-time interaction, *c*_*A*_ is distributed according to the hypergeometric function with mean *µ*_*A*_.

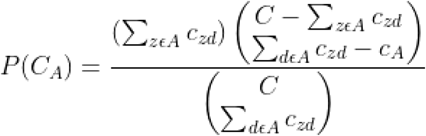

When Σ_*zϵA*_ *c*_*zd*_ and Σ_*dϵA*_ *c*_*zd*_ are small compared to C, *c*_*A*_ is approximately Poisson distributed with mean *µ*_*A*_. Since *c*_*A*_ is approximately Poisson distributed, we can use the Poisson generalized likelihood ratio (GLR) to measure if the scanning window contains an anomalous case count.

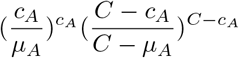

Hypothesis testing is then performed by generating a large number of random permutations of each case in the data set. In this study, we generated 99,999 simulated datasets and evaluated statistical significance using Monte Carlo hypothesis testing. The p-value of each cluster is p = R/(S +1) where R is the rank of the maximum GLR from the real data set and S is the number of simulated data sets. If the GLR from the observed dataset is higher than the 5,000th highest GLM of the simulated dataset, it is considered statistically significant at the 0.05 level.

We conducted three complementary analyses using different levels of spatial aggregation. The most detailed analysis was at the **room-level**-each case was mapped and analyzed by the specific room the infected student lived in at the time of infection (Figure 2). For the second analysis, cases were analyzed at the **floor-level**; all cases on a floor were aggregated to the same location and the location from floor to floor only varied by height (Figure 3). Finally, an **street address-level** scan was conducted, where every case in each building was grouped to a single geographic locus.

**Figure 2.**
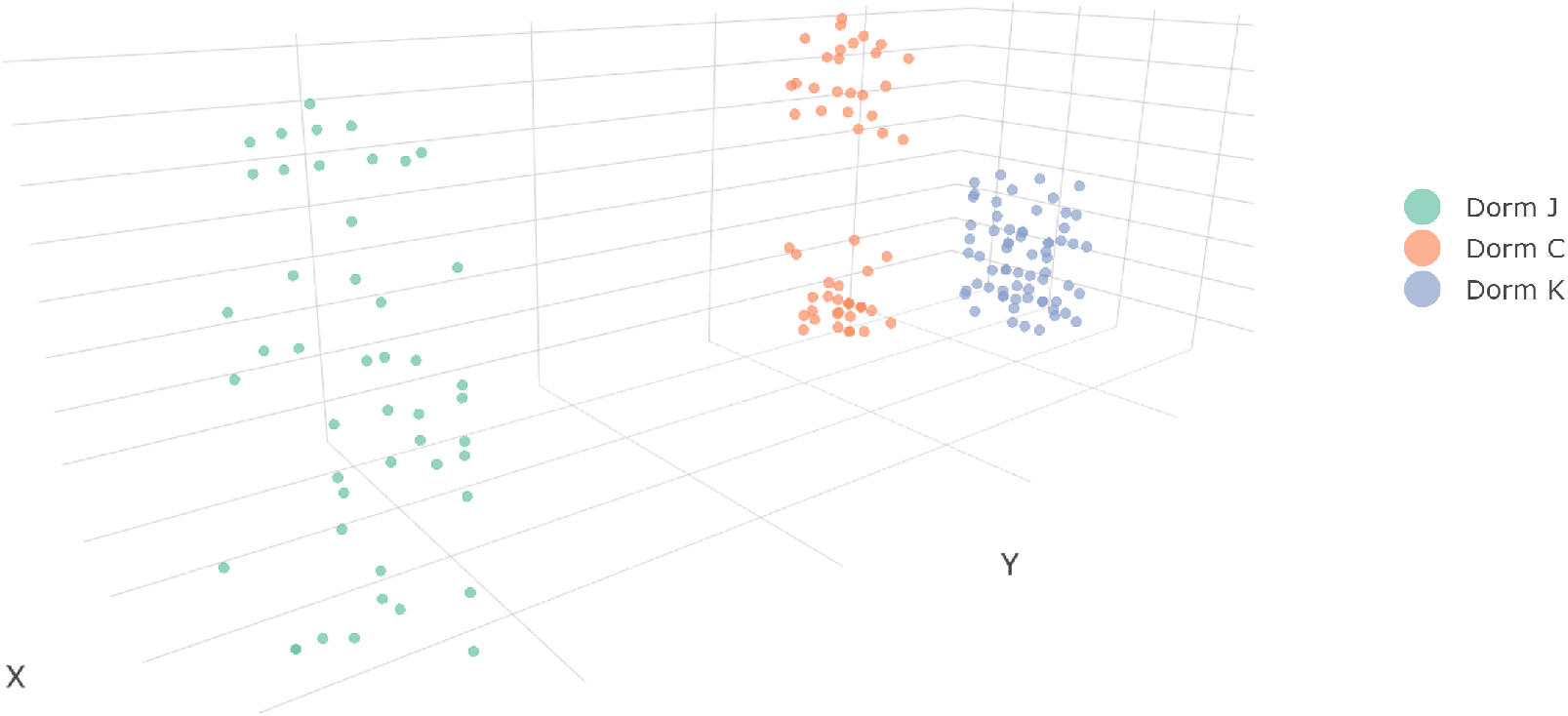
Room-Level 3D Plot of SARS-CoV-2 infections in residence halls, 2021.

**Figure 3.**
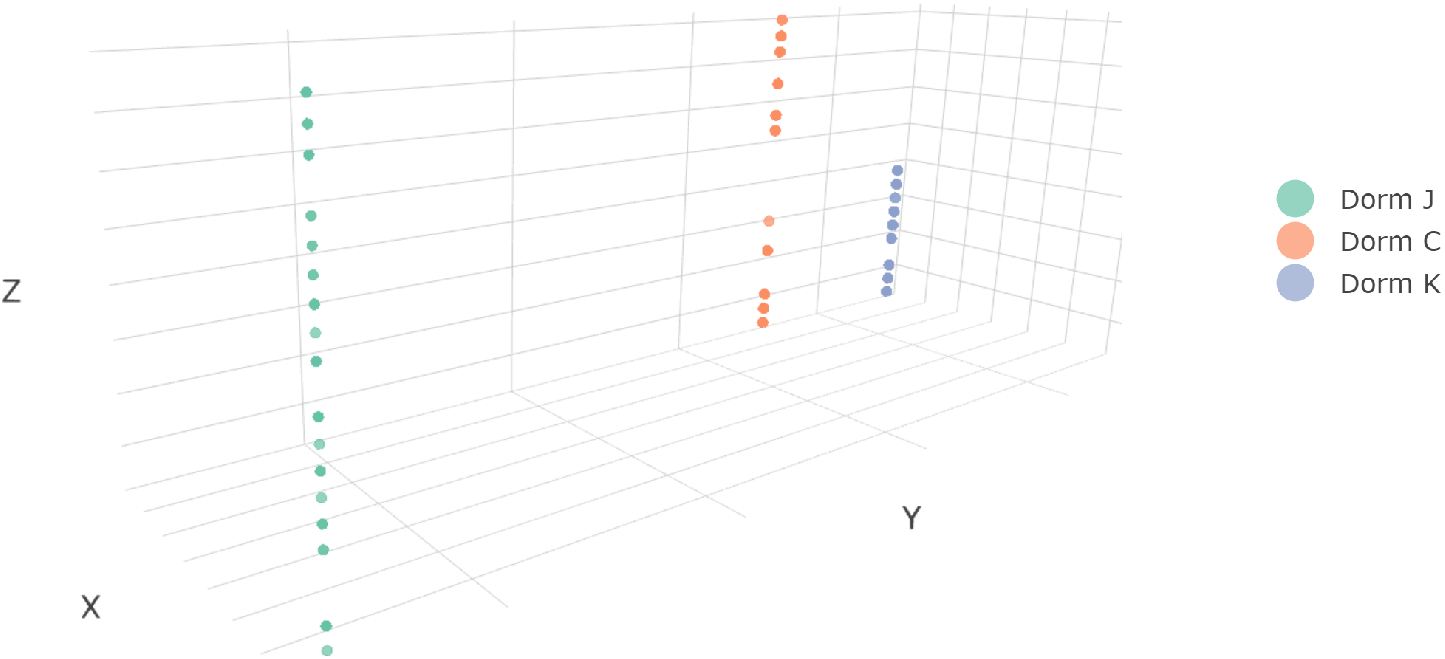
Floor-Level 3D Plot of SARS-CoV-2 infections in residence halls, 2021.

### 2.4 Purely Temporal analysis utilized the CDC Algorithm (EARS)

As a sensitivity analysis for the spatial-temporal scan analyses, we used the CDC Early Aberration Reporting System (“EARS”) as implemented in the R “surveillance” [19] to detect outbreaks in a purely temporal manner. This statistical aberration detection method was chosen because does not require lengthy historical data to be used as a control and instead uses the previous five days to calculate the mean and standard deviation required for the analysis [20]. This method was more appropriate for our analysis as historical data were unavailable due to the recent emergence of COVID-19. While developed for syndromic surveillance (groups of symptoms, as opposed to specific clinical diagnoses) it has been widely used for a range of confirmed infections. We conducted our analysis on the cases of all three residence halls in aggregate, as well as confirmed cases in all three dorms individually. Using a purely temporal algorithm on individual dorms removes the spatial element from this analysis. For these analyses, thresholds were set using a minimum sigma of 1, a 5-day scan window, and with medium sensitivity.

## 3 Results

### 3.1 Room-level analysis

Analysis of case-patients with room-level geolocation yielded three statistically significant clusters. Information about these clusters is contained in table 1. These clusters were contained within Dorm C, save for one case in the third cluster which occurred in Dorm K. The cases in each cluster varied from 5 to 8 different floors with lengths of 7 to 25 days. The spatial overlaps can be seen in Figure 4.

**Table 1:** Room-level clusters of SARS-CoV-2 infections detected using SaTScan (space-time permutation mode), 2021.

| Cluster | Dorm | Start Date | End Date | Duration (Days) | Radius (Meters) | # of Floors | Observed Cases | Expected Cases | Relative Risk | Test Statistic | p-value |
| --- | --- | --- | --- | --- | --- | --- | --- | --- | --- | --- | --- |
| 1 | Dorm C /Dorm K | 15-Feb | 22-Feb | 7 | 36.4 | 6 | 21 | 9.15 | 2.29 | 6.037012 | 0.04464 |
| 2 | Dorm C | 20-Apr | 15-May | 25 | 33.7 | 5 | 10 | 2.11 | 4.73 | 7.841271 | 0.0009 |
| 3 | Dorm C | 5-May | 15-May | 10 | 43.0 | 8 | 9 | 2.14 | 4.21 | 6.211861 | 0.03148 |

**Figure 4.**
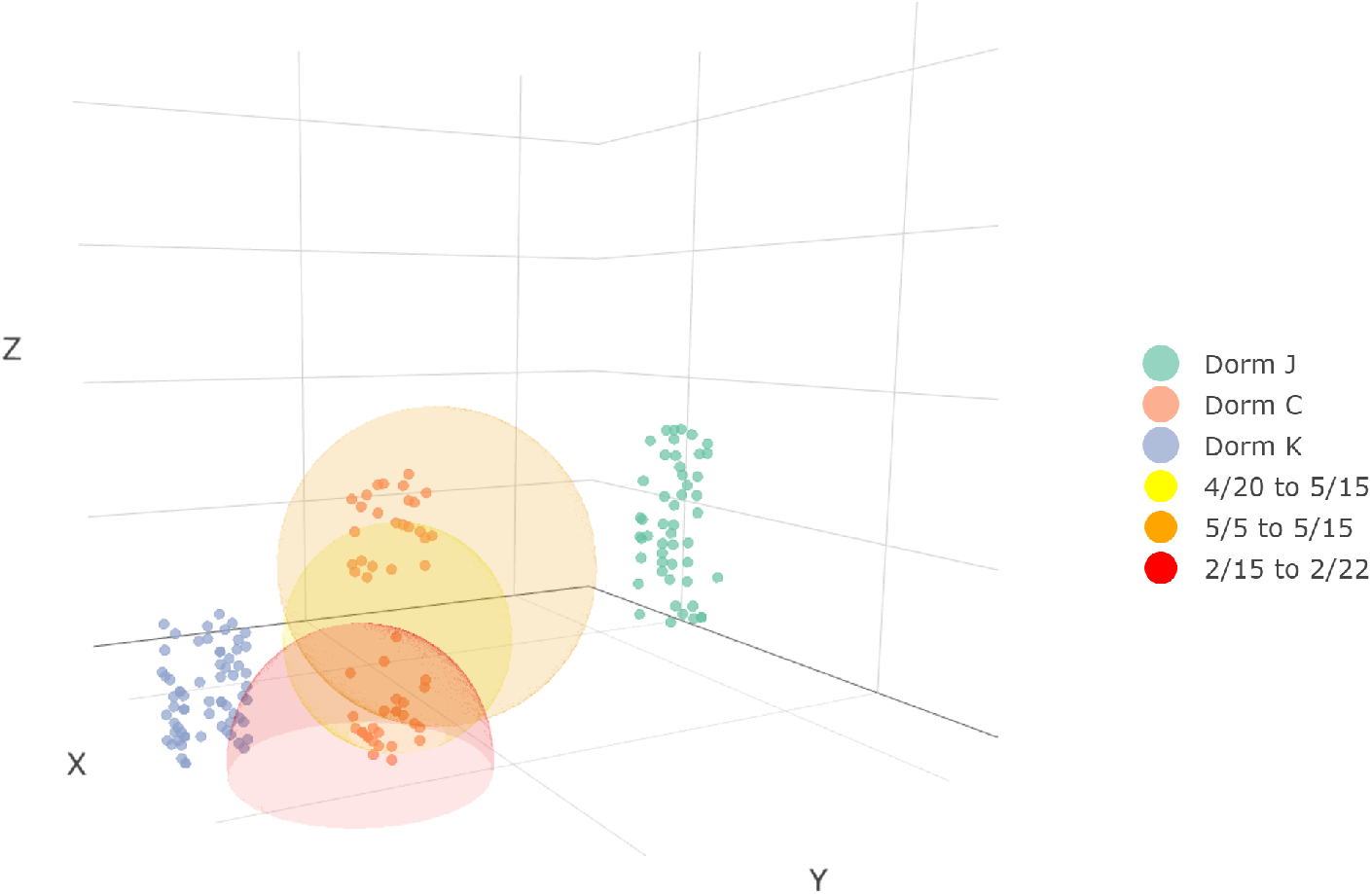
Hotspots of elevated case counts detected using SaTScan, room-level analysis-SARS-CoV-2 in residence halls, 2021. (Note-rotated for clarity.)

### 3.2 Floor-level analysis

Analysis at the floor-level also yielded three statistically significant case-clusters. All of these clusters were identified within Dorm C, and spanned from two to seven floors, with duration of 7 to 25 days. (Table 2).

**Table 2:** Floor-level clusters of SARS-CoV-2 infections detected with SaTScan (space time permutation model), 2021.

| Cluster | Dorm | Start Date | End Date | Duration (Days) | Radius (Meters) | # of Floors | Observed Cases | Expected Cases | Relative Risk | Test Statistic | p-value |
| --- | --- | --- | --- | --- | --- | --- | --- | --- | --- | --- | --- |
| 1 | Dorm C | 15-Feb | 22-Feb | 7 | 7.6 | 2 | 20 | 8.37 | 2.39 | 6.223845 | 0.00248 |
| 2 | Dorm C | 20-Apr | 15-May | 25 | 22.8 | 5 | 10 | 2.16 | 4.62 | 7.657971 | 0.00022 |
| 3 | Dorm C | 5-May | 15-May | 10 | 34.2 | 7 | 9 | 2.14 | 4.21 | 6.211861 | 0.00254 |

### 3.3 Address-level analysis

Two statistically significant address-level clusters were observed, one in Dorm C and one in Dorm J. Both clusters had a reported radius of zero meters due to all cases within a residence all being geolocated to a single point and being completely contained within each dorm. The cluster duration ranged from 25 to 45 days, and contained cases from 11 to 16 floors. (Table 3).

**Table 3:** Address-level clusters of SARS-CoV-2 infections detected using SaTScan’s space time permutation model.

| Cluster | Dorm | Start Date | End Date | Duration (Days) | Radius (Meters) | # of Floors | Observed Cases | Expected Cases | Relative Risk | Test Statistic | p-value |
| --- | --- | --- | --- | --- | --- | --- | --- | --- | --- | --- | --- |
| 1 | Dorm J | 28-Feb | 14-Apr | 45 | 0 | 16 | 14 | 0.41 | 1.89 | 2.454119 | 0.08863 |
| 2 | Dorm C | 20-Apr | 15-May | 25 | 0 | 11 | 11 | 3.47 | 3.17 | 5.341485 | 0.00049 |

### 3.4 CDC EARS

Aggregate cases across all three residence halls, are shown in Figure 5. Retrospective surveillance using the EARS algorithm yielded no time periods with infections above the alarm threshold. Analysis of individual dorms provided an alert for Dorm C on February 15th as shown in Figure 6; an alarm for Dorm K February 25th as shown in figure 7; and no alarms were observed for Dorm J as shown in Figure 8; - supplemental.

**Figure 5.**
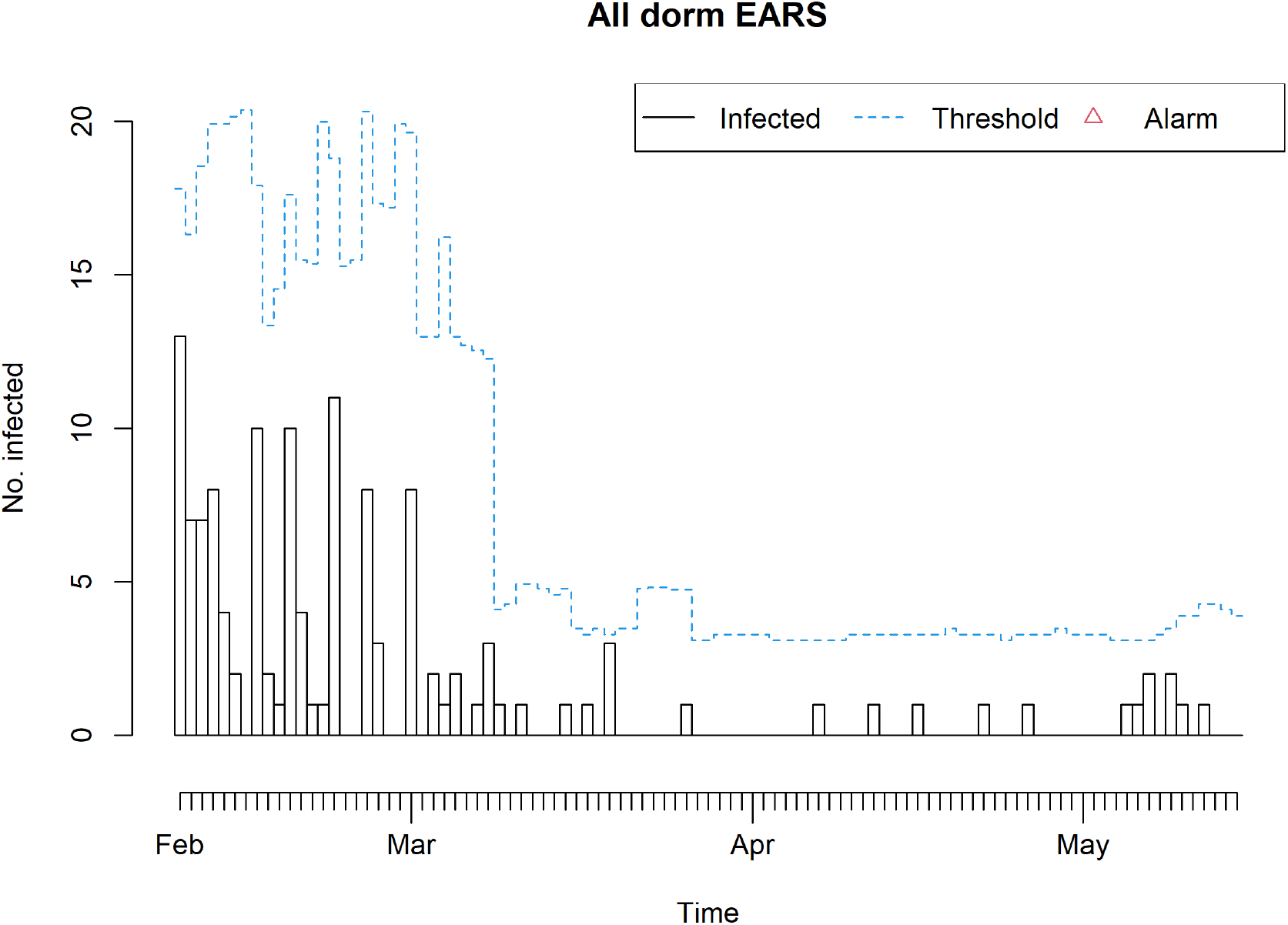
CDC EARS analysis of SARS-CoV-2 cases in residence halls, 2021.

**Figure 6.**
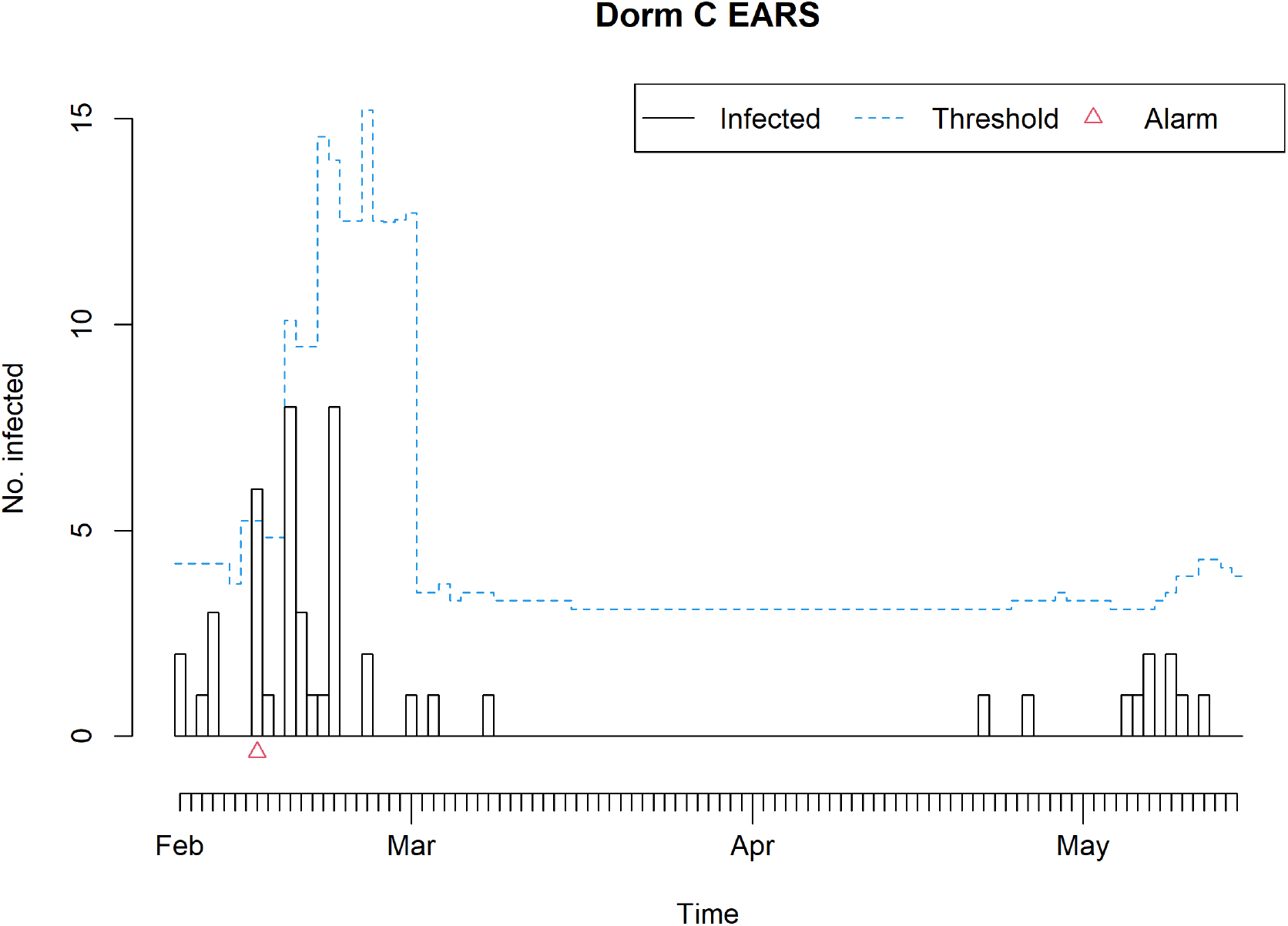
CDC EARS analysis of SARS-CoV-2 cases in Dorm C, 2021.

**Figure 7.**
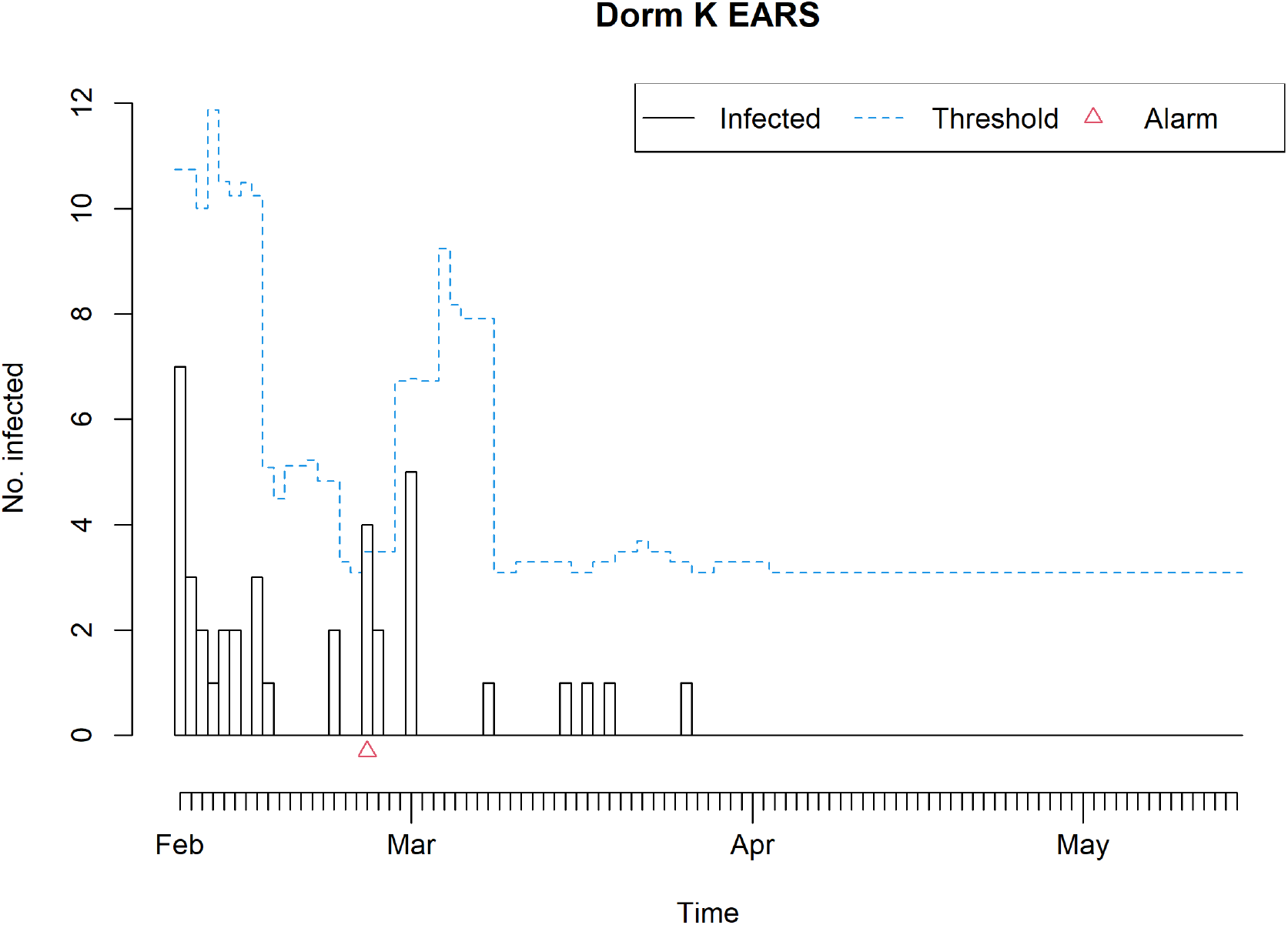
CDC EARS analysis of SARS-CoV-2 cases in Dorm K, 2021.

## 4 Discussion

The goals of retrospective cluster detection are to quantify transmission clusters to gain information to predict future outbreaks, assess possible sources of transmission, and to optimize resource allocations for response programs. Implementing spatial-temporal scan statistics at different spatial scales allows us to consider not only these goals, but also to weigh the effectiveness against the complexity, time, and workload involved with analysis at each level of spatial resolution.

### 4.1 Space-Time Permutation Scan Statistic

Analysis with exact 3-D (room-level) locations provides statistical evidence for three clusters, with cases that were all or mostly within a single residence hall (Dorm C). With analysis at an aggregated floor-level, three statistically significant clusters were also detected, two of which were very consistent with the clusters detected using the room-level data. However, one of these clusters was much smaller in size and included cases on only two floors (as opposed to six in the room-level analysis.

With analysis at the street address-level, there was evidence for two statistically significant clusters, one in the Dorm C and the other in Dorm J. These address-level clusters are smaller in spatial extent than those detected at the other levels of detail, as all the cases in each dorm are centered on a single geographic locus. The first cluster detected at the address level was consistent with the first clusters detected at the floor and room level, with the same start and end dates, but included cases on more floors, with an estimated lower relative risk. The second cluster at the address level not only occurred in a different dorm than those identified by the other detail levels, but also had a duration of 45 days, which is much longer than any other statistically significant clusters identified.

As each model identified at least some statistically significant clusters within the Dorm C, we can conclude that there were probably multiple localized outbreaks within this building over the study period. While the direct causes of these outbreaks are unclear, the clustering of cases strongly suggests that there was local transmission of infection within the Dorm C building itself. In the future, further steps should be taken to prevent the spread of disease in this potentially high-risk setting.

Prior work used simulation studies to examine the the impact of data aggregation on detection of purely spatial clusters (without consideration of temporarily)[21]. This study found near-perfect sensitivity using exact location-data, with decreasing power, and increasing false positive rates, with data aggregation. Moreover, the authors note the false positive rate increases rapidly with the first data aggregation, but comparatively slowly thereafter. However, other researchers report contrary impacts from aggregation with with spatio-temporal algorithms, believed to be due to the very different analyses.[22]

The number of detected cases in Dorms K, C, and J was never large enough to merit a context-specific response. The room-level and floor-level analyses produced comparable results, while the address-level analysis lacked the sensitivity to identify different clusters within the same dorm. Using the maximum number of iterations in the SaTScan program, analysis for the room-level took 54 minutes to complete sufficient iterations. In comparison, the floor-level analysis took just under 5 minutes, and the address-level analysis took 18 seconds. The room-level analysis also required a comparatively complex data processing to geolocate each room, and a much longer data processing time than the floor-level analysis. Moreover, students on the same floor are likely to be epidemiologically linked to one another in this context, as they share restrooms and communal living spaces. The floor-level analysis shows comparable sensitivity with a fraction of the labor and processing, and would appear to be like the most optimal choice for health surveillance in this context.

### 4.2 CDC Early Aberration Reporting System (EARS)

The simpler temporal testing using EARS algorithm detected no statistically significant clustering of cases during the study period when conducted on the case counts from all three dorms in aggregate, presumably as the case count 5-day running means were monotonic and decreasing during this time period. This clear demonstrates the strength of spatio-temporal analyses in health surveillance. Even when appropriately calibrated, a purely temporal analysis has the potential to miss relevant epidemiological signals. When used on the case reporting from individual dorms, EARS did detect elevated risk periods in both Dorm C and Dorm K; however, this analysis does not adjust for multiple testing.

The first date and location flagged by EARS, February 15th in Dorm C, is consistent with the case clusters detected in the floor-level and room-level analyses. The other date and location flagged by EARS (February 25th in Dorm K) does not coincide with any clusters detected in SaTScan analyses; this may be a false-positive.

### 4.3 Limitations

This analysis has several limitations. Foremost, there is no “gold standard” for what constitutes a genuine outbreak in this context; our comparative sensitivity assumes no false-positives in the fully geolocated (room-level) data.

Secondly, the three residence halls studied are near many other dormitories in the region of the UMass campus, and students mixed in many different settings. While this analysis uses student “home address” to reference case-patients, transmission of a respiratory pathogen like SARS-CoV-2 may occur in many other settings unrelated to the residence hall.

Additionally, some of these other residence halls are closer to the three analyzed residence halls than they are to one another. This means that most cross-dorm clustering may be undetected despite being relevant in a public health context. Finally, the Space-Time Permutation model adjusts for purely spatial and temporal clusters, so clusters are detected comparatively, meaning that they are high numbers of cases compared to other locations in the three studied dorms. Unanalyzed transmission in other areas of campus could impact the background rates used for estimating the expected cases.

Another limitation of the study is the implied spatial and epidemiological relationships between reported cases. The analysis was performed in a Euclidean manner, represented in three-dimensional space with the absolute difference in height between the floors also being the distance between cases in height. However, pathogens do not spread through ceilings and floors (generally), nor are students able to move as readily in the vertical (z) direction as they are in the x and y direction along a floor.. The underlying spatial epidemiology between floors in this context may not be properly represented by simple Euclidean space.

Finally, these results may not be representative of case-patient aggregations in routine public health surveillance. University students in general, and especially in the context of COVID disruptions, have unusual contact matrices relative to the general populations[23], Moreover, during the pandemic many major socio-behavioral shifts were documented within the UMass campus community [24].

### 4.4 Recommendations

This study finds that while there are some gains in detections from using exact location data, the “alert” signal is largely unchanged relative to use of floor-level data. That is, sensitivity is comparable, with simpler requirements for geolocation and analysis. However, use of aggregated data (retaining temporarily, but using address-level coordinates) leads to lower sensitivity for alerts.

This analysis would complement other early detection strategies such as purely temporal scans, address-level scans, and concurrent wastewater analysis. Buildings or clusters of buildings with statistically higher case counts or high wastewater viral indicators could be scanned with SaTScan to identify higher-risk floors or groups of floors for targeted public health response.

For future implementations of SaTScan in similar settings, SaTScan’s Poisson model should be considered, which incorporates underlying population data into its analysis. Kulldorff et al report that when reliable population-at risk data are available, the Poisson-based scan statistic model is expected to perform better than the space-time permutation model [18].

Additionally, the use of non-Euclidean methods of modeling the relationship between floors, similar to the model created by Abboud et al used to study the spread of antibiotic resistant *Klebsiella pneumoniae* in a single clinical site [10] should be considered.

## Data Availability

Due to the fine-scale spatiotemporal nature of these data, these data are not publicly available due to patient privacy.

## Revision History

| Revision | Date | Author(s) | Description |
| --- | --- | --- | --- |
| 1.0 | 2021 | KA, AAL | MS thesis |
| 1.1 | 22.11.24 | AAL | Created preprint; edits |
| 1.2 | 27.11.24 | AAL, KA | Updated figures added; edits |
| 1.3 | 19.12.24 | AAL, KA | Updated figures; extensive edits |

## 5 Supplemental figures

**Figure 8.**
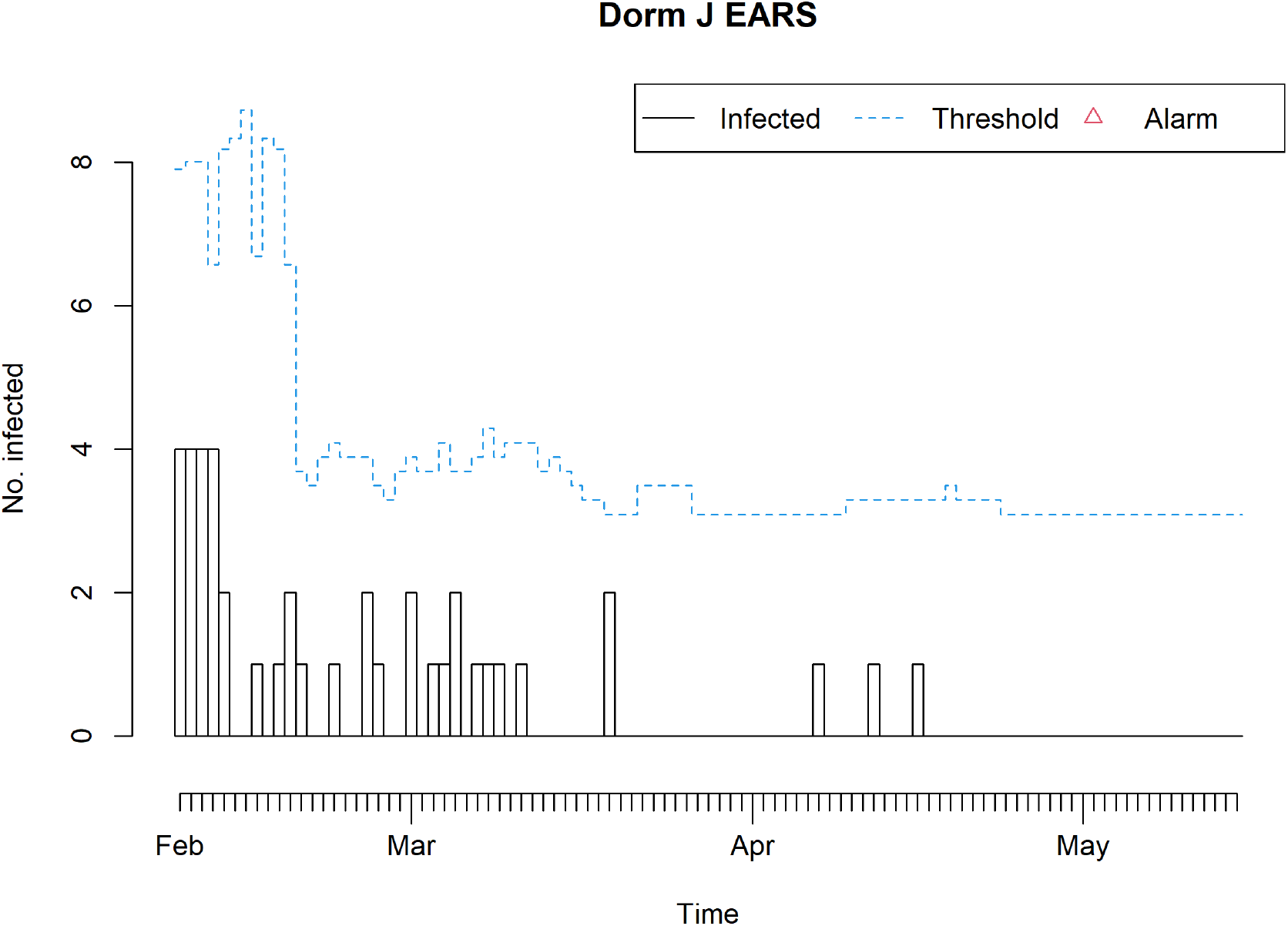
CDC EARS analysis of SARS-CoV-2 cases in Dorm J, 2021.

